# Accelerating cough-based algorithms for pulmonary tuberculosis screening: Results from the CODA TB DREAM Challenge

**DOI:** 10.1101/2024.05.13.24306584

**Authors:** Devan Jaganath, Solveig K Sieberts, Mihaja Raberahona, Sophie Huddart, Larsson Omberg, Rivo Rakotoarivelo, Issa Lyimo, Omar Lweno, Devasahayam J. Christopher, Nguyen Viet Nhung, William Worodria, Charles Yu, Jhih-Yu Chen, Sz-Hau Chen, Tsai-Min Chen, Chih-Han Huang, Kuei-Lin Huang, Filip Mulier, Daniel Rafter, Edward S.C. Shih, Yu Tsao, Hsuan-Kai Wang, Chih-Hsun Wu, Christine Bachman, Stephen Burkot, Puneet Dewan, Sourabh Kulhare, Peter M. Small, Vijay Yadav, Simon Grandjean Lapierre, Grant Theron, Adithya Cattamanchi

**Affiliations:** Division of Pediatric Infectious Diseases, University of California, San Francisco, San Francisco, USA; Center for Tuberculosis, University of California, San Francisco, USA; Sage Bionetworks, Seattle, USA; CHU Joseph Rasera Befelatanana, Antananarivo, 101, Analamanga, Madagascar; Centre d’Infectiologie Charles Mérieux, Université d’Antananarivo, Antananarivo, 101, Analamanga, Madagascar; Department of Epidemiology and Biostatistics, University of California, San Francisco, San Francisco, CA, USA; CHU Tambohobe Fianarantsoa, 301, Haute-Matsiatra, Madagascar; Université de Fianarantsoa, Fianarantsoa, 301, Haute-Matsiatra, Madagascar; Ifakara Health Institute, Environmental and Ecological Sciences & Interventions and Clinical Trials Departments, Kiko Avenue, Plot 463, Mikocheni, Dar es Salaam, Tanzania; Christian Medical College, Vellore (Ranipet campus), Tamil Nadu, India; National Tuberculosis Programme, 463 Hoang Hoa Tham, Ba Dinh District, Hanoi, Vietnam; VNU, University of Medicine and Pharmacy; Walimu, Plot 341 White Close Najjera – Wakiso district, Kampala, Uganda; De La Salle Medical and Health Sciences Institute, Governor D. Mangubat Avenue, Dasmarinas Cavite, Philippines 4114; Graduate Institute of Biomedical Electronics and Bioinformatics, National Taiwan University, Taipei, Taiwan; Industrial Information Department, Development Center for Biotechnology, Taipei, Taiwan; Investment & Wealth Management, FCC Partners Inc., Taipei, Taiwan; Graduate Program of Data Science, National Taiwan University and Academia Sinica, Taipei, Taiwan; Research Center for Information Technology Innovation, Academia Sinica, Taipei, Taiwan; Department of Data Science, ANIWARE, Taipei, Taiwan; School of Medicine, China Medical University, Taichung, Taiwan; Flywheel.io, Minneapolis, Minnesota USA; Institute of Biomedical Sciences, Academia Sinica, Taipei, Taiwan; Independent Researcher, Taipei, Taiwan; Artificial Intelligence and E-learning Center, National Chengchi University, Taipei, Taiwan; Global Health Labs, 14360 SE Eastgate Way, Bellevue, USA; Department of Global Health, University of Washington, Seattle, USA; Hyfe, Inc, Seattle, USA; Centre de Recherche du Centre Hospitalier de l’Université de Montréal, Immunopathology Axis, 900 St-Denis, Montréal, Québec, H2X 0A9 Canada; Université de Montréal, Department of Microbiology, Infectious Diseases and Immunology, 2900 Edouard-Montpetit, Montréal, Québec, H3T 1J4 Canada; DSI-NRF Centre of Excellence for Biomedical Tuberculosis Research, South African Medical Research Council Centre for Tuberculosis Research, Division of Molecular Biology and Human Genetics, Faculty of Medicine and Health Sciences, Stellenbosch University, South Africa; University of California Irvine, School of Medicine, Orange, USA; Department of Computer Science, Ashoka University, Haryana, India; Klick Applied Sciences, Klick Inc., 175 Bloor Street East, Toronto, Canada; Department of Computational Biology, Indraprastha Institute of Information Technology, New Delhi, India; AI campus, Cedar Sinai, Los Angeles, CA, USA; Department of Computational Medicine & Bioinformatics, University of Michigan - Ann Arbor. Ann Arbor, MI, USA; Arkansas AI Campus, AR, USA; Xi’an Jiaotong-Liverpool University, Wisdom Lake Academy of Pharmacy, Suzhou, China; Cancer Data Science Laboratory, National Cancer Institute, National Institutes of Health, Bethesda, Maryland, USA; Bentonville High School, Bentonville, Arkansas, USA

## Abstract

**Importance:** Open-access data challenges have the potential to accelerate innovation in artificial-intelligence (AI)-based tools for global health. A specimen-free rapid triage method for TB is a global health priority.

**Objective:** To develop and validate cough sound-based AI algorithms for tuberculosis (TB) through the Cough Diagnostic Algorithm for Tuberculosis (CODA TB) DREAM challenge.

**Design:** In this diagnostic study, participating teams were provided cough-sound and clinical and demographic data. They were asked to develop AI models over a four-month period, and then submit the algorithms for independent validation.

**Setting:** Data was collected using smartphones from outpatient clinics in India, Madagascar, the Philippines, South Africa, Tanzania, Uganda, and Vietnam.

**Participants:** We included data from 2,143 adults who were consecutively enrolled with at least two weeks of cough. Data were randomly split evenly into training and test partitions.

**Exposures:** Standard TB evaluation was completed, including Xpert MTB/RIF Ultra and culture. At least three solicited coughs were recorded using the Hyfe Research app.

**Main Outcomes and Measures:** We invited teams to develop models using 1) cough sound features only and/or 2) cough sound features with routinely available clinical data to classify microbiologically confirmed TB disease. Models were ranked by area under the receiver operating characteristic curve (AUROC) and partial AUROC (pAUROC) to achieve at least 80% sensitivity and 60% specificity.

**Results:** Eleven cough models were submitted, as well as six cough-plus-clinical models. AUROCs for cough models ranged from 0.69-0.74, and the highest performing model achieved 55.5% specificity (95% CI 47.7-64.2) at 80% sensitivity. The addition of clinical data improved AUROCs (range 0.78-0.83), five of the six submitted models reached the target pAUROC, and highest performing model had 73.8% (95% CI 60.8-80.0) specificity at 80% sensitivity. In post-challenge subgroup analyses, AUROCs varied by country, and was higher among males and HIV-negative individuals. The probability of TB classification correlated with Xpert Ultra semi-quantitative levels.

**Conclusions and Relevance:** In a short period, new and independently validated cough-based TB algorithms were developed through an open-source and transparent process. Open-access data challenges can rapidly advance and improve AI-based tools for global health.

**Key Points:** *Question:* Can an open-access data challenge support the rapid development of cough-based artificial intelligence (AI) algorithms to screen for tuberculosis (TB)?

*Findings:* In this diagnostic study, teams were provided well-characterized cough sound data from seven countries, and developed and submitted AI models for independent validation. Multiple models that combined clinical and cough data achieved the target accuracy of at least 80% sensitivity and 60% specificity to classify microbiologically-confirmed TB.

*Meaning:* Cough-based AI models have promise to support point-of-care TB screening, and open-access data challenges can accelerate the development of AI-based tools for global health.

## Introduction

As global health challenges intersect with rapid advancements in technology and artificial intelligence (AI), digital health tools have the potential to enhance disease surveillance, diagnosis, and management.^1–3^ In particular, the widespread availability of smartphones and wearable sensors create opportunities for low-cost, non-invasive applications to increase healthcare access and quality.^4^ However, development and deployment of AI solutions have primarily focused on commercial applications in high-income markets. A major challenge to equitable implementation of these tools is a lack of available datasets from diverse geographic settings, and limited focus on conditions that disproportionally affect low- and middle-income countries (LMICs).^2,5^ Moreover, available datasets may be proprietary, preventing open-access sharing and transparent algorithm development. The consequence is a dearth of AI tools validated in LMICs and that address the public health challenges they face.

Tuberculosis (TB) is the leading cause of death from an infectious disease worldwide.^6^ The high mortality is driven by a large case detection gap, in which 3.1 million of the estimated 10 million individuals who develop TB disease each year have not been diagnosed or reported to public health programs.^6^ AI has already supported TB diagnosis through automated chest X-ray reading, and computer-assisted detection algorithms (CAD) have been endorsed by the World Health Organization (WHO) as a triage tool.^7^ However, CAD systems require infrastructure and expertise to obtain chest X-rays which limit their impact at primary health facility levels. Cough is a common symptom of TB, and initial studies suggest that there are unique acoustic features that can distinguish pulmonary TB from other respiratory conditions.^8,9^ Furthermore, cough detection applications have already been developed for mobile phones and smart watches,^10^ providing an opportunity to integrate cough-based AI algorithms for point-of-care TB assessment by providers and patients.

In other diseases, including COVID-19,^11^ open-access, crowd-sourced data challenge initiatives have been used to accelerate the development of novel algorithms.^12^ These initiatives provide a transparent platform to share methods and findings, and support independent validation. To expedite AI diagnostic development for TB, we established a cough sound repository from individuals prospectively enrolled with presumptive TB across seven high TB-burden countries, and launched the Cough Diagnostic Algorithm for TB (CODA TB) Dream Challenge.^13^ We present the results of the challenge and highlight the role of this approach to rapidly advance AI tools for global diseases that impact LMICs.

## Methods

### CODA TB DREAM Challenge

The CODA TB DREAM Challenge launched on October 26, 2022. Participants were asked to develop a model to classify TB disease in two sub-challenges: (1) using cough sounds alone and (2) using cough sounds and basic demographic and clinical variables. Challenge teams were allowed to submit results for one or both sub-challenges. To be considered in the official ranking, teams needed to submit a final report that outlined their methods and conclusions, and a link to their source code. The timeline of the challenge is shown in **Supplemental Figure 1**.

The challenge was hosted by Sage Bionetworks, which has developed an open-science, collaborative competition framework for evaluating and comparing computational algorithms, using the DREAM Challenges framework. DREAM focuses exclusively on biomedicine with an explicit mandate for transparency, openness, and collaboration. The challenge was set up on Synapse (www.synapse.org/tbcough), which provided all instructions, a secure platform for data sharing, a forum for communication with challenge participants, and supported submission of models for independent validation.

Any individual or team could participate in the challenge. After registering for a free Synapse account, they certified that they understood the Synapse data use policy, verified their identify, and agreed to the challenge guidelines to not attempt to identify or contact any study participants, to not share the data with others, and that they must comply with the intended use of the data. If they agreed to these conditions, they were given access to the de-identified training data as described below. The challenge was advertised as broadly as possible, including on social media, to multiple academic institution listservs and departments of global health, bioinformatics and computer science, companies interested in cough-based or TB diagnosis, and previous DREAM Challenge participants.

### Study dataset

Data for the CODA TB DREAM Challenge were obtained from two multi-country TB diagnostic evaluation studies.^13^ The Rapid Research in Diagnostic Development TB Network (R2D2 TB Network) enrolled participants at outpatient health centers in Uganda, South Africa, Vietnam, the Philippines and India. The Digital Cough Monitoring Project enrolled participants in Tanzania and Madagascar. Ethical approvals for the studies were obtained from institutional review boards (IRB) in the US (R2D2 TB Network, University of California, San Francisco) and Canada (Digital Cough Monitoring Project, University of Montreal), as well as IRBs in each country in which participants were enrolled. All participants provided written informed consent for study participation, cough recording and anonymized data sharing.

In both studies, eligible participants were 18 years or older and had a new or worsening cough for at least two weeks. Participants completed a standard evaluation for TB including a clinical questionnaire and examination, sputum-based molecular testing (Xpert MTB/RIF Ultra, Cepheid, Sunnyvale) and liquid or solid medium culture testing. Participants were asked to produce at least three solicited cough sounds during the baseline visit prior to any TB treatment initiation. The coughs were collected on an Android-based smartphone using the Hyfe Research app,^14^ which uses a convolutional neural network (CNN) model to automatically detect the cough and saves the 0.5 second peak sound.^13^ Solicited cough sounds were collected, though any triggered passive coughs were also recorded. TB disease status was based on a microbiological reference standard, defined by a positive molecular or culture result. Further details on the study procedures and dataset including a summary of participant demographics and country distribution have been published previously.^13^

A training set (n=1,105) for algorithm development was created by taking a 50% sample of the dataset randomized at the individual level. Of the remaining data, 24% (n=248) was randomly selected at the individual level for the “leaderboard” test set, from which challenge participants could receive periodic feedback on their model performance, and the remainder (n=790) was reserved as the final test set for algorithm evaluation. Challenge teams were given direct access only to the training set, which included the raw peak cough sound recordings in WAV format, as well as associated age, sex, height, weight, smoking status, self-reported duration of cough, history of prior TB, common TB symptoms (hemoptysis, fever, night sweats, weight loss), heart rate and temperature. These variables were chosen as data that would be readily available in routine primary care settings. We did not include HIV status as the testing and/or results may not be available or known at the time of cough assessment.

### Algorithm development and evaluation

Participating teams could train an algorithm using any pre-processing approach and model, and with any programming language (i.e. R, Python, etc.) or framework (such as Keras or Pytorch). For evaluation, models were required to be saved in Open Neural Network Exchange (ONNX) format and submitted in a Docker container, with any code needed for pre-processing the data.

Challenge teams had five interim opportunities to evaluate their algorithms on the “leaderboard” test set before the final algorithms were due for test set evaluation (**Supplemental Figure 1**). The teams submitted their preliminary models and we independently applied those models to the leaderboard test set. The output of each model was continuous TB prediction scores used to generate calculate area under the receiver operating characteristic curve (AUROC) and two-way partial AUROC^15^ (pAUROC) with 80% sensitivity and 60% specificity. We set this threshold to identify promising algorithms that had an accuracy that was at least within 10% of the minimum WHO target product profile (TPP) accuracy for a TB triage test (≥90% sensitivity, ≥70% specificity).^16^ A higher pAUROC indicates that a greater area meets the minimum target sensitivity and specificity. Original model evaluation was performed in Python (version 3.8.8). All subsequent analyses were performed in R Software version 4.2.2 (2022-10-31). Evaluations of model statistics were done using the pROC R package. Implementation of the pAUROC was provided by Chaibub Neto, et al.^17,18^

The final submission deadline was on February 13, 2023, four months from the launch of the challenge. Similar to the leaderboard rounds, challenge teams submitted their pre-processing code (if applicable) and trained models, and we independently applied the model to the test set. Final model performance was evaluated by the pAUROC. If no model could meet the accuracy threshold, the algorithms were evaluated by the total AUROC. Variability in the AUROCs and pAUROCs were assessed via bootstrap resampling (n = 1,000).

### Clinical Data Only Model

As a sensitivity analysis to assess the degree that the clinical variables alone contributed to the models in sub-challenge 2, we developed a Random Forest model^19^ using the clinical and demographic variables provided to challenge participants. In addition to the variables provided, body mass index (BMI) was computed from height and weight variables, and duration of cough symptoms was log-transformed prior to model fitting. The model was trained using 1,000 trees.

### Subgroup analyses

After the challenge was complete, first- and second-ranked teams (n=5 due to ties) in both sub-challenges were invited to participate in additional model evaluation. We assessed the accuracy of the models by country, sex, and HIV status. We also compared the probability of TB classification for each model by Xpert MTB/RIF Ultra semi-quantitative PCR result.

## Results

### Challenge Implementation

147 individuals and 18 teams registered for the CODA TB DREAM Challenge. In each leaderboard round, two to eight teams submitted models. Thirteen teams submitted final models for sub-challenge 1, and eight for sub-challenge 2. Of those that submitted final models, eleven (sub-challenge 1) and six (sub-challenge 2) teams submitted a summary of methods and model code. The winning models for each sub-challenge are described in the Supplemental Methods. The reports for the full set of submissions are available through the challenge website.^20^

### Sub-challenge 1

As shown in **Table 1**, **Figure 1A** and **Supplemental Figure 2**, AUROCs ranged from 0.689 to 0.743. The top model achieved a specificity of 55.5% at 80% sensitivity; as further shown in **Supplemental Table 1**, it did not achieve the WHO TPP-based accuracy thresholds for a triage test at 90% sensitivity and 70% specificity. Of the 11 groups, 4 (36%) used CNNs, 4 (36%) used artificial neural networks, and 3 (27%) used gradient boosting decision tree methods.

**Figure 1.**
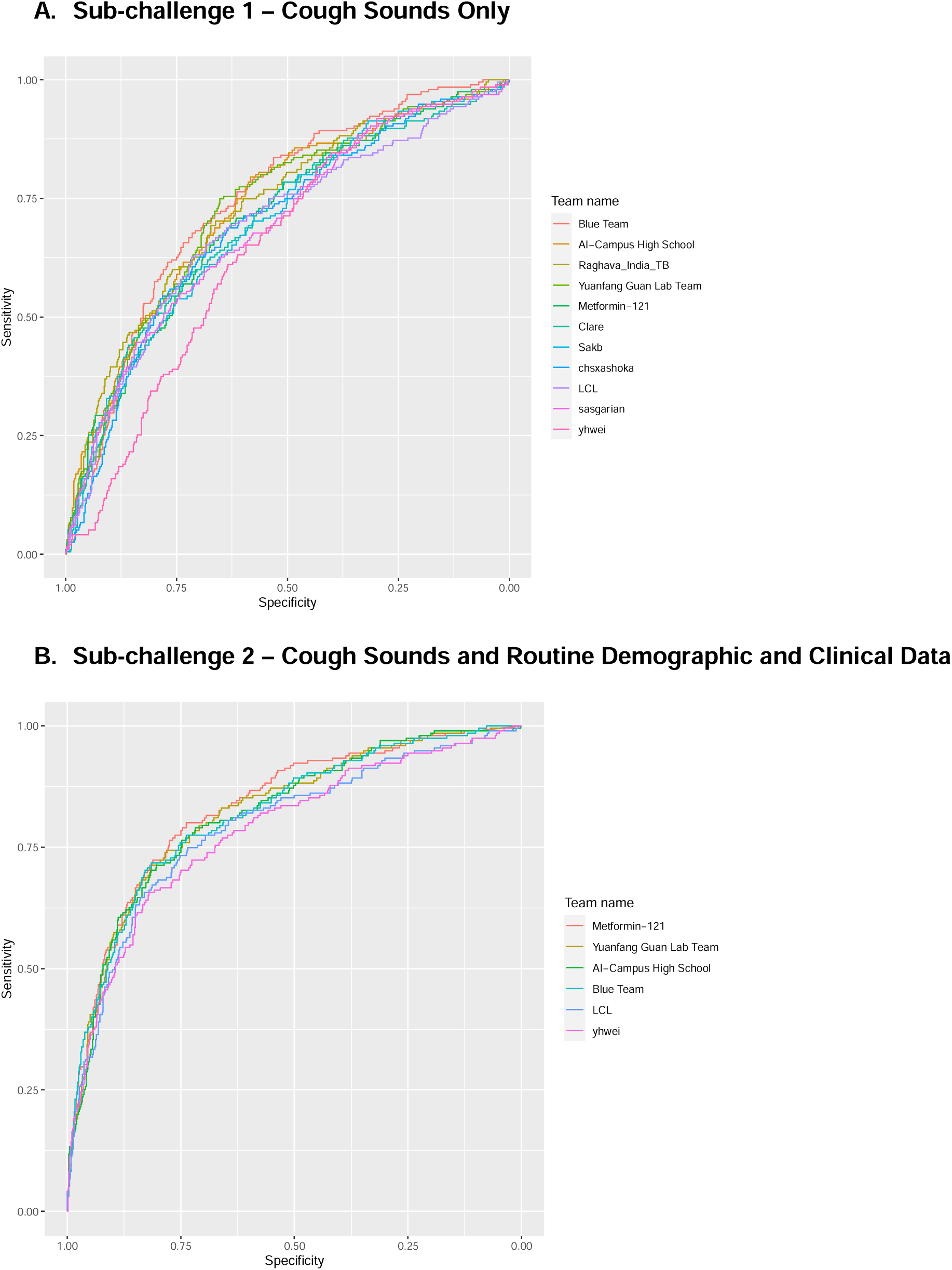
Receiver Operating Characteristic Curves for CODA TB DREAM Challenge Final Models.

**Table 1.**
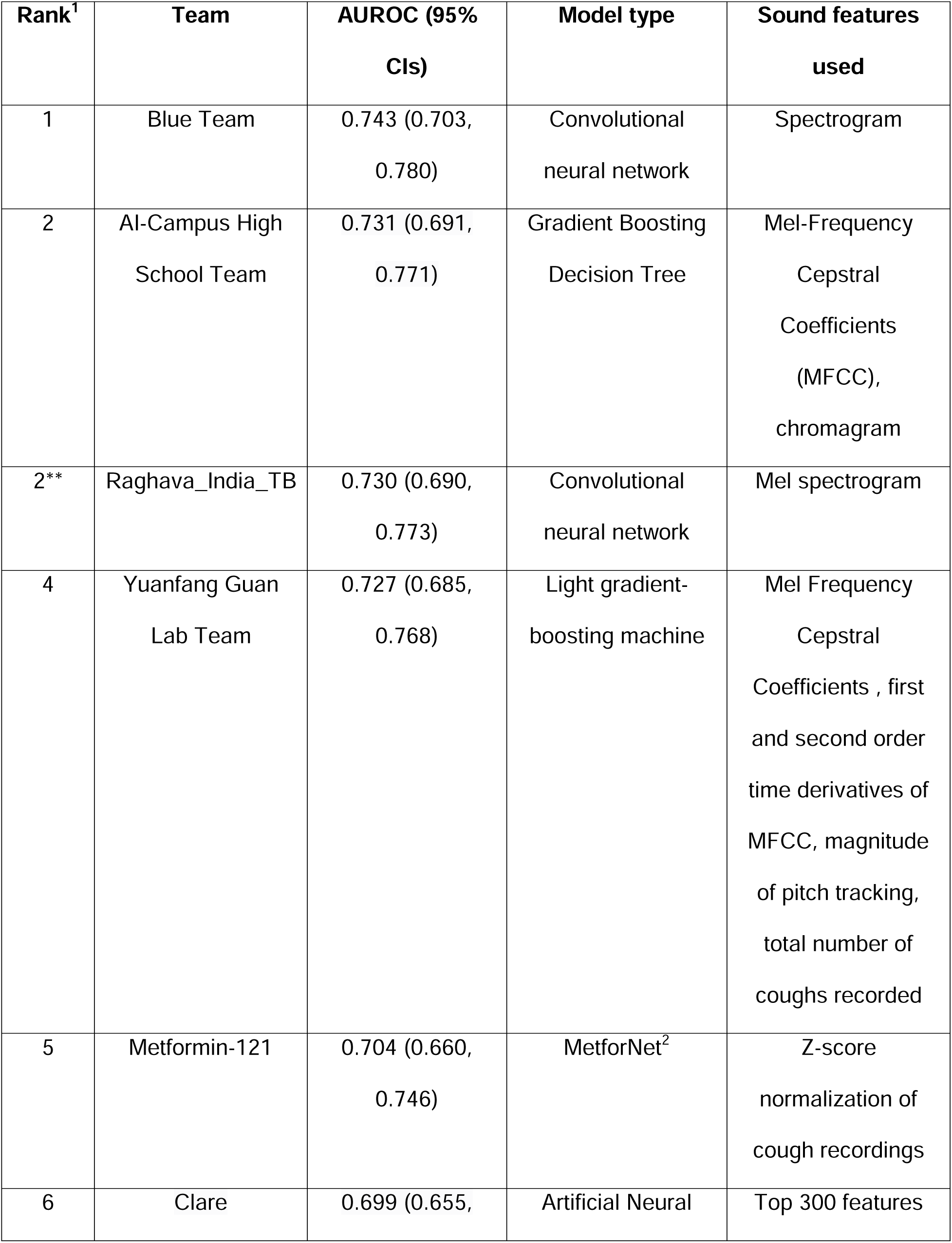
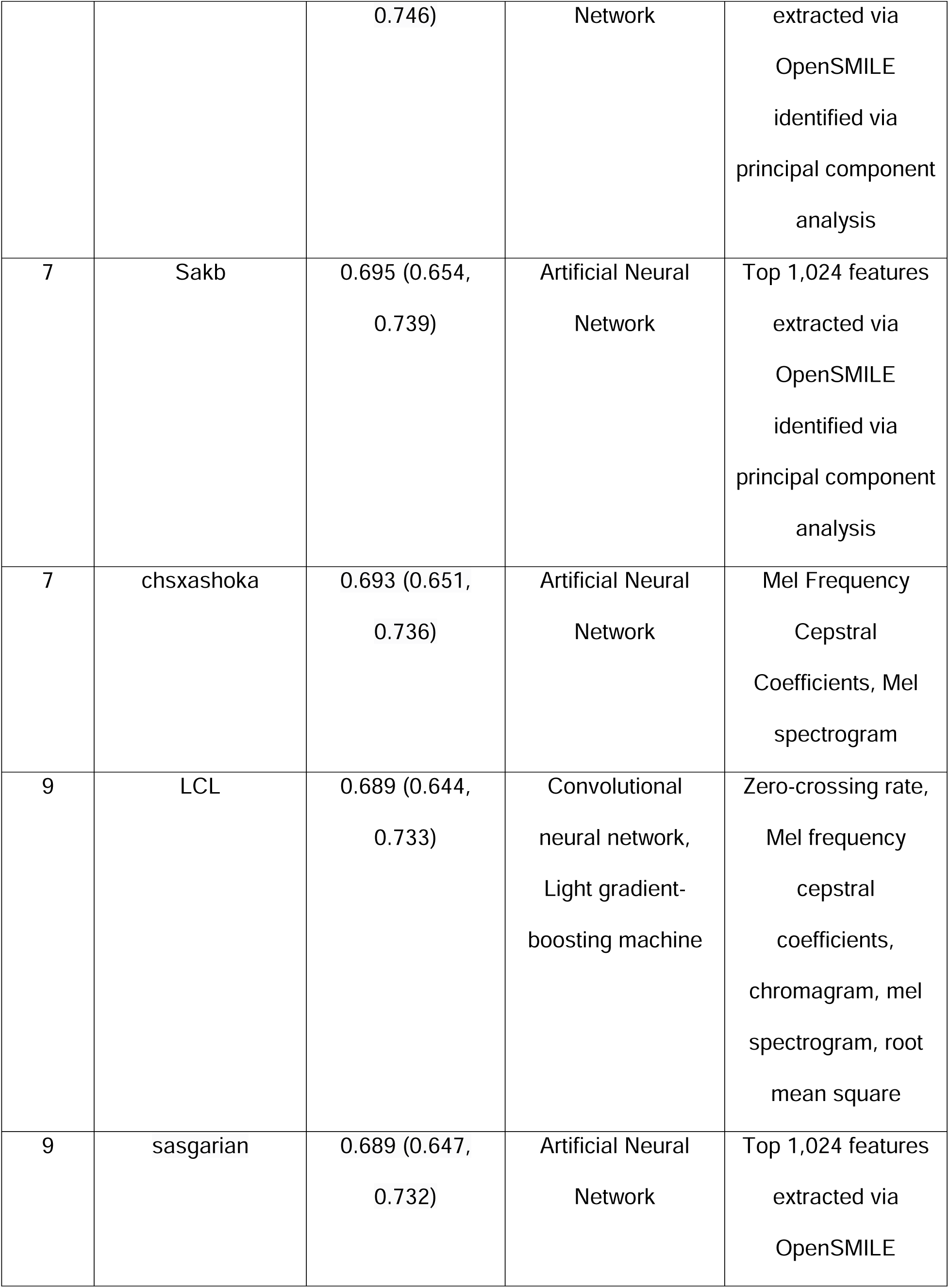
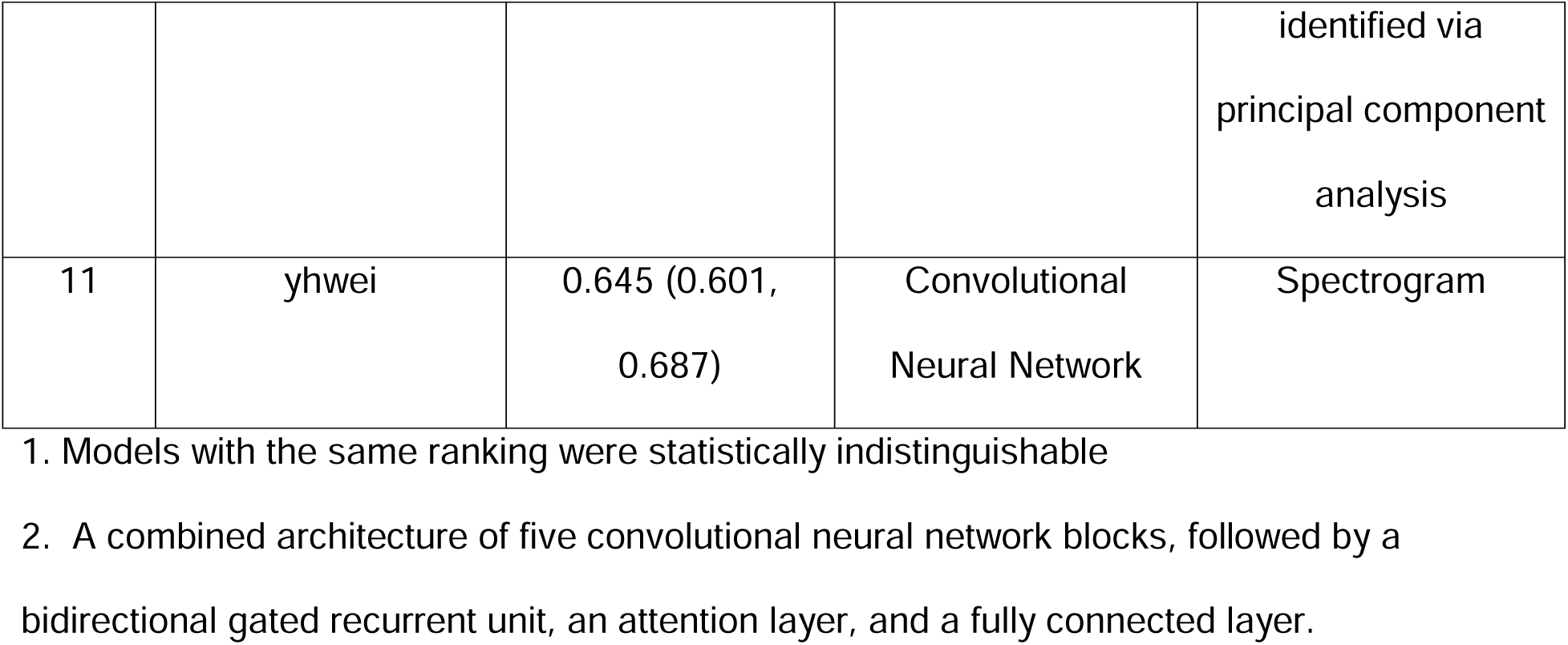
Model performance for cough-only model (Sub-challenge 1)

### Sub-challenge 2

All groups used the same algorithm approach they utilized in sub-challenge 1. As shown in **Figure 1B, Supplemental Figure 3** and **Table 2**, overall performance improved compared to the use of cough sounds alone, and the top performing model achieved an AUROC of 0.832 (95% CI 0.795-0.863) and a pAUROC of 0.003 (95% CI 6.1e-06-0.012). Five of the six (83%) submission achieved at least 80% sensitivity and 60% specificity, with the top model reaching 73.8% (95% CI 60.8-80.0) at 80% sensitivity. For the WHO TPP for a TB triage test, the top performing model achieved 54% specificity (95% CI = (38%, 63%)) at 90% sensitivity (**Supplemental Table 1**). In sensitivity analysis, the clinical data only model achieved an AUROC of 0.817 (95% CI 0.778-0.850) and pAUROC of 0.004 (95% CI 5.5e-4-0.010). This was higher than the cough only model, but the top combined cough and clinical data model outperformed both.

**Table 2.** Model performance for cough sound and clinical data model (Sub-challenge 2)

| Rank | Team | pAUROC (95% CI) | AUROC (95% CI) |
| --- | --- | --- | --- |
| 1 | Metformin-121 | 0.003 (6.11e-06, 0.012) | 0.832 (0.795, 0.863) |
| 2 | Yuanfang Guan Lab Team | 0.003 (0, 0.009) | 0.821 (0.784, 0.853) |
| 3 | AI-Campus High School<br>Team | 0.001 (0, 0.008) | 0.817 (0.778, 0.850) |
| 4 | Blue Team | 0.001 (0, 0.007) | 0.818 (0.779, 0.853) |
| 5 | LCL | 0.001 (0, 0.006) | 0.792 (0.750, 0.829) |
| 6 | yhwei | 0 (0, 0.003) | 0.784 (0.741, 0.822) |

### Subgroup Assessment

Model performance for the combined cough sound and clinical data models (sub-challenge 2) was variable across country of data collection (**Figure 2A**). In general, the models performed better on data from the Philippines, Uganda, Tanzania and Vietnam. The median AUROC of the cough and clinical data models was slightly higher for males compared to females (median 0.82 vs. 0.78, p<0.01, **Figure 2B**). Model performance was also slightly higher among people not living with HIV compared to people living with HIV (median AUROC 0.83 vs. 0.78, p<0.01, **Figure 2C**). Subgroup results for sub-challenge 1 (cough sounds only) are shown in **Supplemental Figures 4-7**. Findings were similar, although we found slightly lower accuracy in males vs. females (median AUROC 0.69 vs 0.71, p = 0.02) in contrast to sub-challenge 2.

**Figure 2.**
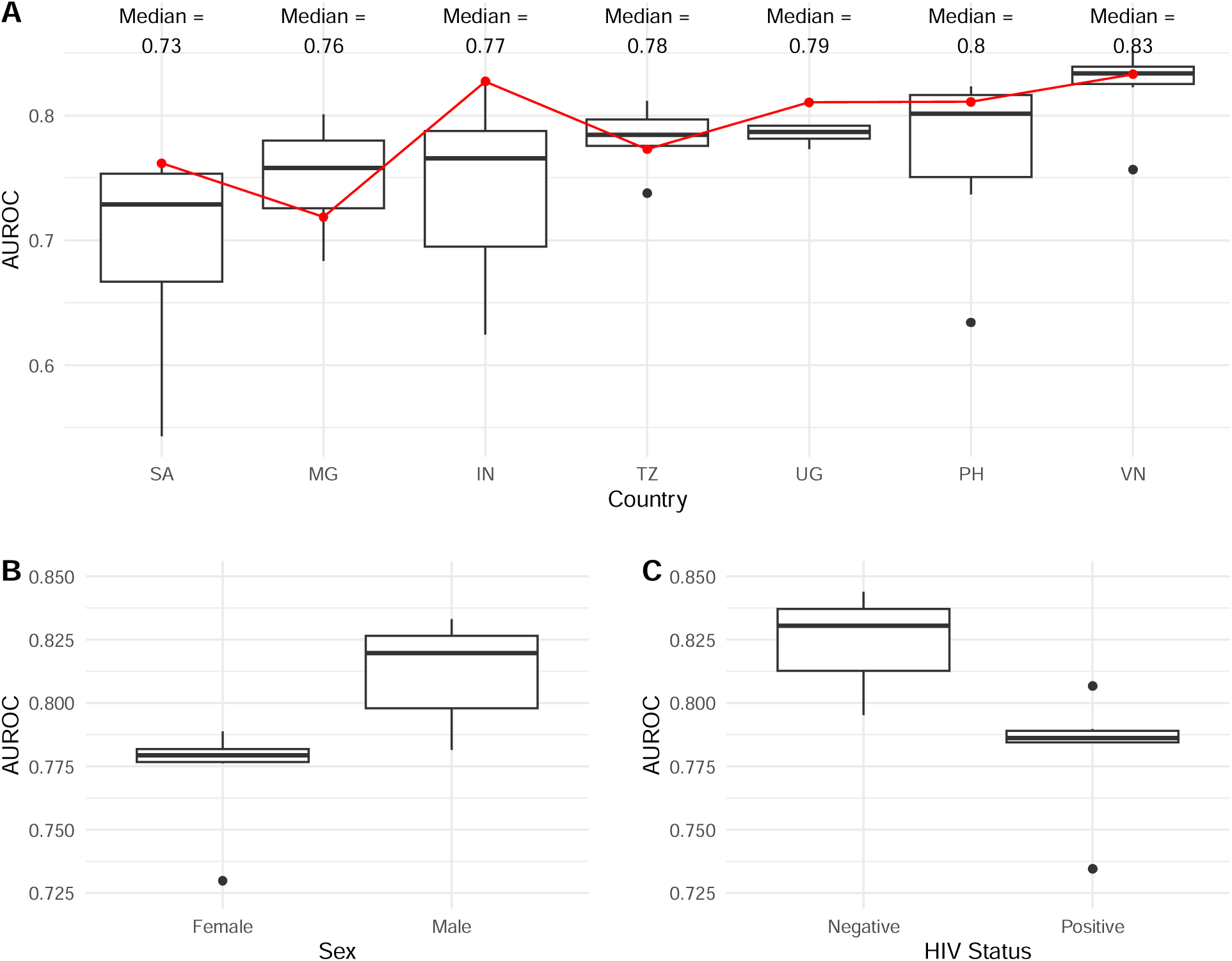
Comparison of Area under the Receiver Operating Characteristic Curve by Country and Subgroup in Sub-challenge 2. Box plots of median area under the curve (AUROC) with interquartile range (IQR) based on all submissions, and stratified by (A) country; (B) sex and (C) HIV status. For (A), median AUROC indicated at the top, and winning model AUROC shown in red. SA: South Africa; MG: Madagascar; IN: India; TZ: Tanzania; UG: Uganda; PH: Philippines; VN: Vietnam.

For all submitted cough and clinical data models, the median predicted probability of being TB-positive increased with Xpert MTB/RIF Ultra semi-quantitative level, from trace positive results to high bacillary load results (**Figure 3, Supplemental Figure 7**).

**Figure 3.**
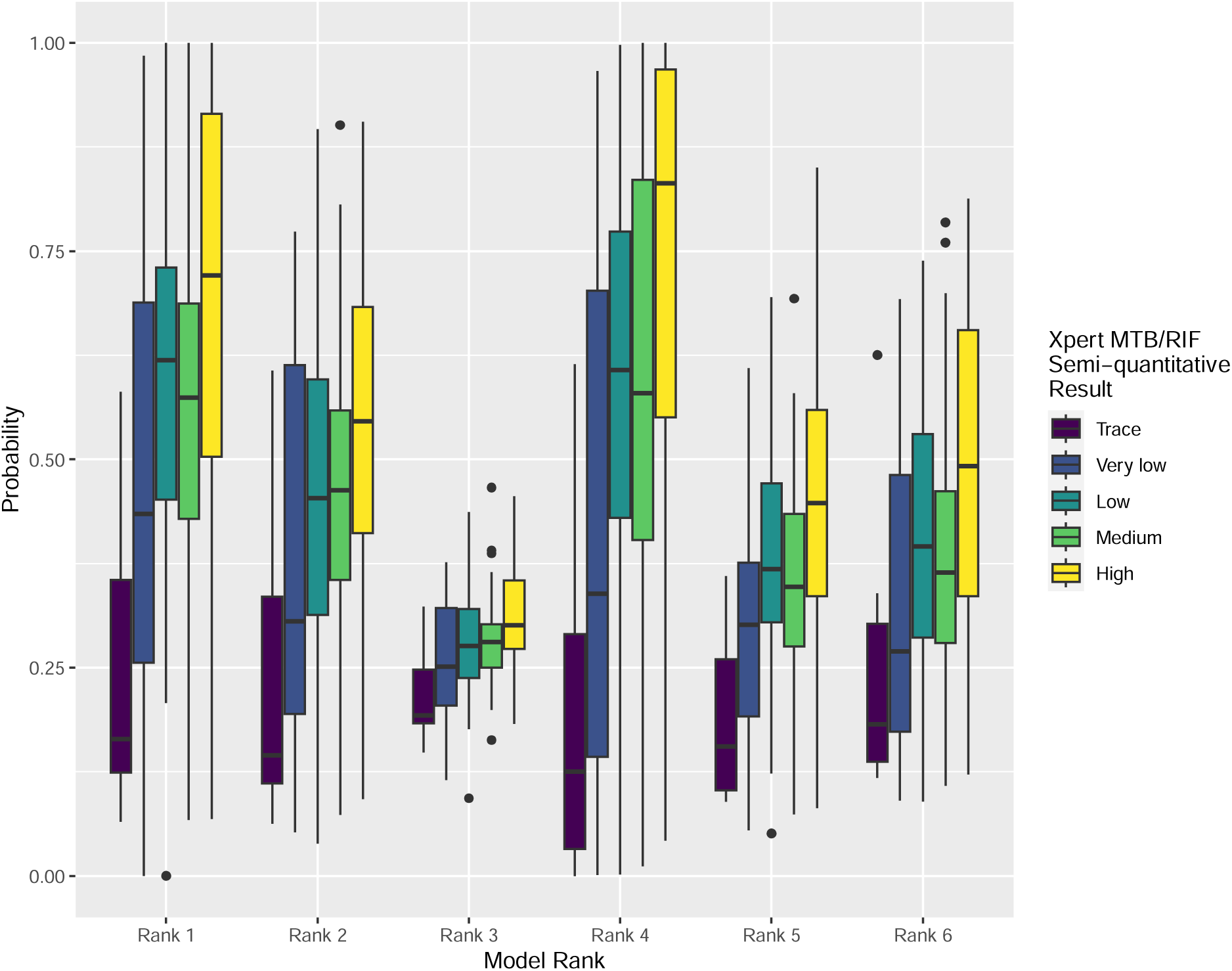
TB probability scores of the cough sound and clinical data models stratified by Xpert semi-quantitative status. Box plot of the median probability with interquartile range (IQR). Rank indicates the final challenge ranking. Higher probability scores indicate higher likelihood that the model would classify the individual as having TB.

## Discussion

The CODA TB Dream Challenge addressed a critical need to accelerate the development of AI-based tools for global health through an inclusive, open and transparent approach. The challenge brought together students, researchers, and industry partners from a diverse geographical spectrum with a common goal of developing novel TB diagnostic algorithms using cough sounds. In a short period, challenge participants created, tested and improved algorithms using cough sounds and routine clinical and demographic data that approached the WHO TPP accuracy targets for a TB triage test. Open-access research and citizen science represent a potential paradigm shift in how digital health solutions can be developed for global health by harnessing the collective expertise of an international community to address a common scientific and humanitarian goal.

The cough-sound only models had similar accuracies with AUROCs that ranged from 0.65-0.74. This performance is within the wide range of cough-based models that have been developed to detect COVID-19 (AUROCs 0.62 to 0.98).^21–24^ In India, for example, a cough-based CNN model achieved an AUROC of 0.75 to detect COVID-19.^24^ There are a few published cough-sound models in TB that have shown higher performance (AUROC 0.79-0.94),^8,9^ but these have been small studies that were not validated on independent datasets, and may overestimate accuracy. A major limitation of previous cough models for other conditions was the use of crowd-sourced data.^11,25,26^ While this approach rapidly generates large real-world datasets, there are multiple challenges, including selection bias, subjective clinical assessment and heterogenous reference standard definitions. In CODA, we utilized a multi-country cohort of consecutively enrolled symptomatic individuals with indications for TB evaluation, standardized clinical data and cough collection protocols, objective TB testing and uniform case definitions. This increases the confidence that algorithms are identifying features specific to the disease condition, reduces AI-related biases, and better reflects how the algorithms will perform in the intended settings and populations.

Performance improved when routine demographic and clinical variables were added to models, and five of six algorithms approached the WHO-established target accuracy thresholds for a TB triage test. We chose demographic and clinical variables that are associated with TB and could be collected in primary care settings or self-reported on a mobile application. As a post-challenge sensitivity analysis, we developed a clinical data only model that performed well (AUROC 0.817), but the addition of cough sound data improved accuracy and supports the role of integrating both data types. The best performing challenge models utilized deep learning algorithms; while interpretability can be limited with such models, subgroup findings increase confidence in a TB-specific signal. First, the probability of TB classification correlated with bacterial burden as measured by semi-quantitative PCR results in both sub-challenges, which was also seen in a recent study in Kenya.^8^ Moreover, worse performance among people living with HIV who often have paucibacillary disease may be expected.^27^ These differences in accuracy have also been seen in CAD algorithms for chest x-ray interpretation,^28^ and different thresholds may be needed depending on the setting or target group.^29^

It is important to recognize that the final submitted models were developed rapidly over a short timeframe, and there is potential for further optimization. This includes exploring more complex CNN architectures and/or ensembles, increasing the size of the training set and developing country-specific models. At the same time, the overarching goal of the challenge was to accelerate innovation and gain key insights into cough-based AI models for TB. In four months, the challenge 1) supported multiple new and independently validated cough sound algorithms that could discriminate TB disease; 2) demonstrated that clinical data could augment performance; and 3) transparently shared the best performing algorithms and processing methods.

To further facilitate ongoing model development, the dataset remains open-source and can be downloaded at the challenge website.^30^ Moreover, the website supports continuous benchmarking so that developers can submit their algorithms to receive independent feedback on model performance. Through this iterative process, the goal is to support the development of at least one cough-based algorithm that could be integrated into a simple mobile device and provide a point-of-care TB triage tool which could be deployed in community-based settings. Once developed, the continuous benchmarking mechanism and held-out data could potentially support its review by a regulatory body.

The dataset and challenge had some limitations. The cough sounds collected were restricted to 0.5 second recordings around the peak; the use of whole cough sounds may further improve performance.^31^ As all participants were symptomatic, there are limitations in extending these models for community-wide screening, and additional data collection from screening cohorts is needed. The participants also all had cough; while solicited cough sounds may have value for those without cough, this needs to be further evaluated. Variation by country may reflect differences in co-morbidities and disease presentation, but also may be due to differences in phone model used and environmental noise. However, 0.5 second recordings limited background noise and algorithms should be developed to be compatible with multiple phone models and environments. The goal of the challenge was to classify microbiologically-confirmed TB; if these algorithms are used as part of two-step screening to guide further testing, other outcomes could be considered such as a radiographic evidence of lung disease. The greater probability of TB classification in individuals with higher bacillary loads may be a useful marker of infectiousness and needs further study. By establishing the platform and approach, additional challenges can be created that update datasets and goals to support new algorithms.

In conclusion, the CODA TB Dream Challenge accelerated the development of cough-sound models that can be integrated into mobile devices for a simple, point-of-care triage tool for TB. It also highlighted how open science and collaborative efforts can support rapid, inclusive, and impactful health innovations. Through such initiatives, we move closer to realizing the expansive potential of digital tools for TB and global health.

## Data Sharing

The challenge training data and links to the code and write-ups for the model submissions are available at www.synapse.org/TBcough. Additionally, users can register to submit models for evaluation against the validation data in an ongoing manner.

## Acknowledgements

The CODA TB DREAM Challenge and post-challenge evaluation was funded in part by the Bill & Melinda Gates Foundation. R2D2 was funded by the U.S. National Institutes of Health (U01 AI152087), and the Digital Cough Monitoring study was funded by the Patrick J. McGovern Foundation. SGL is supported by a Junior 1 Salary Award from the Fonds de Recherche Santé Québec. DJ is supported by funding by the National Institutes of Health. GT acknowledges funding from the EDCTP2 programme supported by the European Union (RIA2018D-2509, PreFIT; RIA2018D-2493, SeroSelectTB; RIA2020I-3305, CAGE-TB) and the National Institutes of Health (D43TW010350; U01AI152087; U54EB027049; R01AI136894).

## Author Contributions

Author contributions include conception (SKS, LO,CB, PD, PMS, SGL, AC), data acquisition (MR, RR, IL, OL, DJC, NVN, WW, CY, GT), data analysis (SKS, SH, JYC, SHC, TMC, CHH, KLH, FM, DR, ESCS, YT, HKW, CHW, SB, SK, VY, and consortium), data interpretation (DJ, SKS, SH, SB, SK, VY, SGL, AC), drafting the manuscript (DJ, SKS, SH), and reviewing the manuscript critically for important intellectual content and final approval of the version to be published (all co-authors).

We agree to be accountable for all aspects of the work in ensuring that questions related to the accuracy or integrity of any part of the work are appropriately investigated and resolved (DJ, SKS, MR, SGL, GT, AC).

