## Supplemental for "Accelerating cough-based algorithms for pulmonary tuberculosis screening: Results from the CODA TB DREAM Challenge"

### Supplemental Methods

**Winning Method (Cough Only)**

Convolutional neural networks applied to the magnitude of the short-term Fourier transform (STFT), also known as spectrogram, have shown promising results for classifying a wide range of sounds. Typically, the mel transformation is used on the frequency bands to more closely match the frequency resolution of the human auditory system. However, for the classification of cough pathology, experiments have demonstrated improved results using the frequency bands without a mel transformation. The cough recordings naturally exhibit a wide variation over a relatively small data set of 1082 subjects. The scoring metric for this challenge is area under the ROC curve (AUROC). In order to improve our estimate of the AUROC metric, 5 fold cross-validation was used for model validation versus a simple training/validation split.

A resnet34 network was used for the majority of trials after showing improved score over resnet18 on initial trials. Pre-trained resnet34 weights were used for initialization. A dropout-rate of 50% was used. The optimization was using Adam with an initial learning rate of 0.0005 and 30 epochs. A one-cycle learning rate schedule was used. The training process involved random resampling of training data to ensure balanced class distributions were presented to the learning algorithm as raw data had a participant class imbalance of 808:297 (roughly 3:1). Processing took roughly 30 min per epoch, or a total of 75 hours for 5 cross-validation folds of 30 epochs on a RTX3090 GPU with a batch size of 64.

**Winning Method (Cough + Metadata)**

The best model incorporating clinical metadata utilized MetforNet, a general end-to-end network solution to predict patterns from different types of biomedical signals which has been previously described by the team members of Metformin-121 [1-4]. This model was utilized as the backbone model to predict the patterns of tuberculosis (TB). The structure comprises a composite architectural framework featuring five convolutional neural network (CNN) blocks, succeeded by a bidirectional gated recurrent unit (GRU), an attention layer [5,6], and a fully connected layer. In architectural design, each CNN block is composed of two convolutional layers, followed by a subsequent convolutional-pooling layer, which is a pooling layer based on convolutional operations. All convolution layers employ a uniform kernel configuration, featuring 12 kernels, each with a kernel size of three. However, an exception exists in the case of the five consecutive convolution-pooling layers, which are distinguished by varying kernel sizes of 24, 24, 24, 24, and 48, in that specific order. In addition, a dropout technique was implemented at a rate of 20% for the connections within the CNN blocks and other independent layers, such as the connections between the last CNN block and the bidirectional GRU layer. This dropout process was introduced to enhance network generalization. Furthermore, batch normalization was incorporated to normalize and rescale the outputs originating from the attention layer. This particular layer employs a specialized mechanism aimed at creating importance-weighting vectors [7]. The Leaky Rectified Linear Unit (LeakyReLU) activation functions were employed in all layers except for the fully connected layer, which featured the utilization of a sigmoid activation function [8]. This architectural framework was developed utilizing the Keras library, with support from TensorFlow running on Graphics Processing Units (GPUs) [9].

To utilize the additional demographic and clinical data, the categorical and numerical data were processed by one-hot encoding and Z-score normalization, respectively. The processed additional demographic and clinical data then concatenate with the output vector of the attention layer above to go through another path of fully connected, batch normalization, LeakyReLU, and dropout layers to produce an additive value by a fully connected layer with a hyperbolic tangent activation function on the output value of the sigmoid activation function to generate another model output. The cough sound records were subjected to Z-score normalization. The dataset was randomly partitioned into 10 equal subsets to establish an 8-1-1 distribution for the purposes of machine learning, encompassing training, validation, and testing phases. Within this data segmentation, the model underwent a training process spanning 100 epochs, representing complete cycles of feeding the training dataset, resulting in the generation of 100 distinct models, each corresponding to a single epoch. The model demonstrating the highest performance on the validation set was subsequently designated as the optimal model for this training iteration. Following this, the superior model was employed to compute the area under the complete Receiver Operating Characteristic (ROC) curve score on the test set. This procedure was iterated 10 times to complete a 10-fold training process, leading to the selection of the top-performing model for each of the 10 folds. The training procedure was executed utilizing the ADAM optimizer in conjunction with a mean-square error loss function [10]. Finally, the 10 best models were merged into a model ensemble by simply averaging their output probability. For the patients with more than one cough sound record, each record was predicted by the model ensemble separately. Still, only the prediction with the highest TB positive probability was selected to make the final decision for each patient.

**Supplemental Table 1. Specificity at 80% and 90% sensitivity for each CODA TB DREAM Challenge submission**

| | Team | Specificity at 80% Sensitivity (95% CI) | Specificity at 90% Sensitivity (95% CI) |
|---|---|---|---|
| **Rank**[1] | Sub-challenge 1 | | |
| 1 | Blue Team | 0.555 (0.466, 0.640) | 0.350 (0.254, 0.487) |
| 2 | AI-Campus High School | 0.571 (0.472, 0.629) | 0.299 (0.207, 0.481) |
| 2 | Raghava_India_TB | 0.504 (0.407, 0.612) | 0.343 (0.208, 0.422) |
| 4 | Yuanfang Guan Lab Team | 0.560 (0.427, 0.661) | 0.289 (0.224, 0.415) |
| 5 | Metformin-121 | 0.476 (0.387, 0.558) | 0.281 (0.180, 0.390) |
| 6 | Clare | 0.469 (0.376, 0.561) | 0.235 (0.141, 0.398) |
| 7 | Sakb | 0.474 (0.363, 0.526) | 0.323 (0.222, 0.383) |
| 7 | chsxashoka | 0.437 (0.336, 0.533) | 0.277 (0.192, 0.345) |
| 9 | LCL | 0.412 (0.287, 0.548) | 0.193 (0.126, 0.299) |
| 9 | sasgarian | 0.429 (0.356, 0.509) | 0.313 (0.198, 0.370) |
| 11 | yhwei | 0.435 (0.368, 0.496) | 0.299 (0.210, 0.382) |
| | Sub-challenge 2 | | |
| 1 | Metformin-121 | 0.738 (0.608, 0.800) | 0.539 (0.403, 0.624) |
| 2 | Yuanfang Guan Lab Team | 0.694 (0.597, 0.782) | 0.442 (0.358, 0.603) |
| 3 | AI-Campus High School | 0.689 (0.548, 0.775) | 0.455 (0.340, 0.556) |
| 4 | Blue Team | 0.657 (0.538, 0.773) | 0.472 (0.339, 0.553) |
| 5 | LCL | 0.645 (0.511, 0.745) | 0.351 (0.267, 0.513) |
| 6 | yhwei | 0.602 (0.454, 0.692) | 0.388 (0.244, 0.459) |

1. Final challenge ranking by AUROC/pAUROC

**Supplemental Figure 1. Timeline of the CODA TB Dream Challenge**

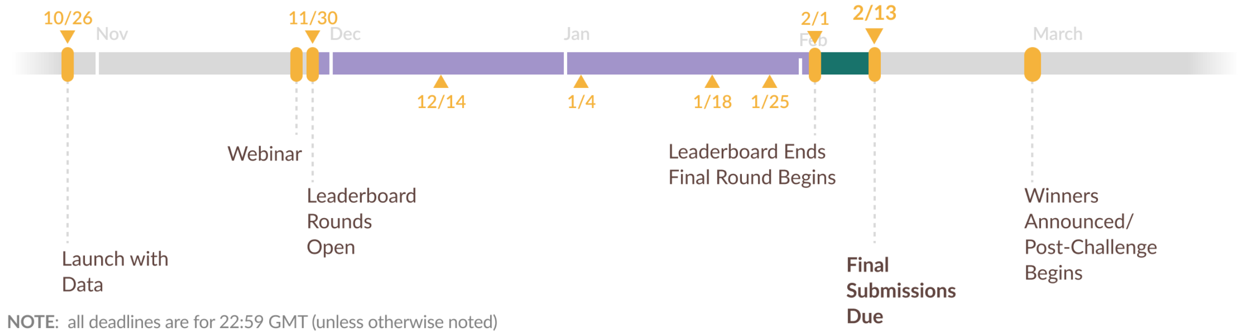

**Supplemental Figure 2. Comparison of AUROCs of submitted sub-challenge 1 (cough data only) models.** Box plot of the median area under the ROC curve (AUROC) of each team with interquartile range (IQR) after 1,000 bootstraps**.**

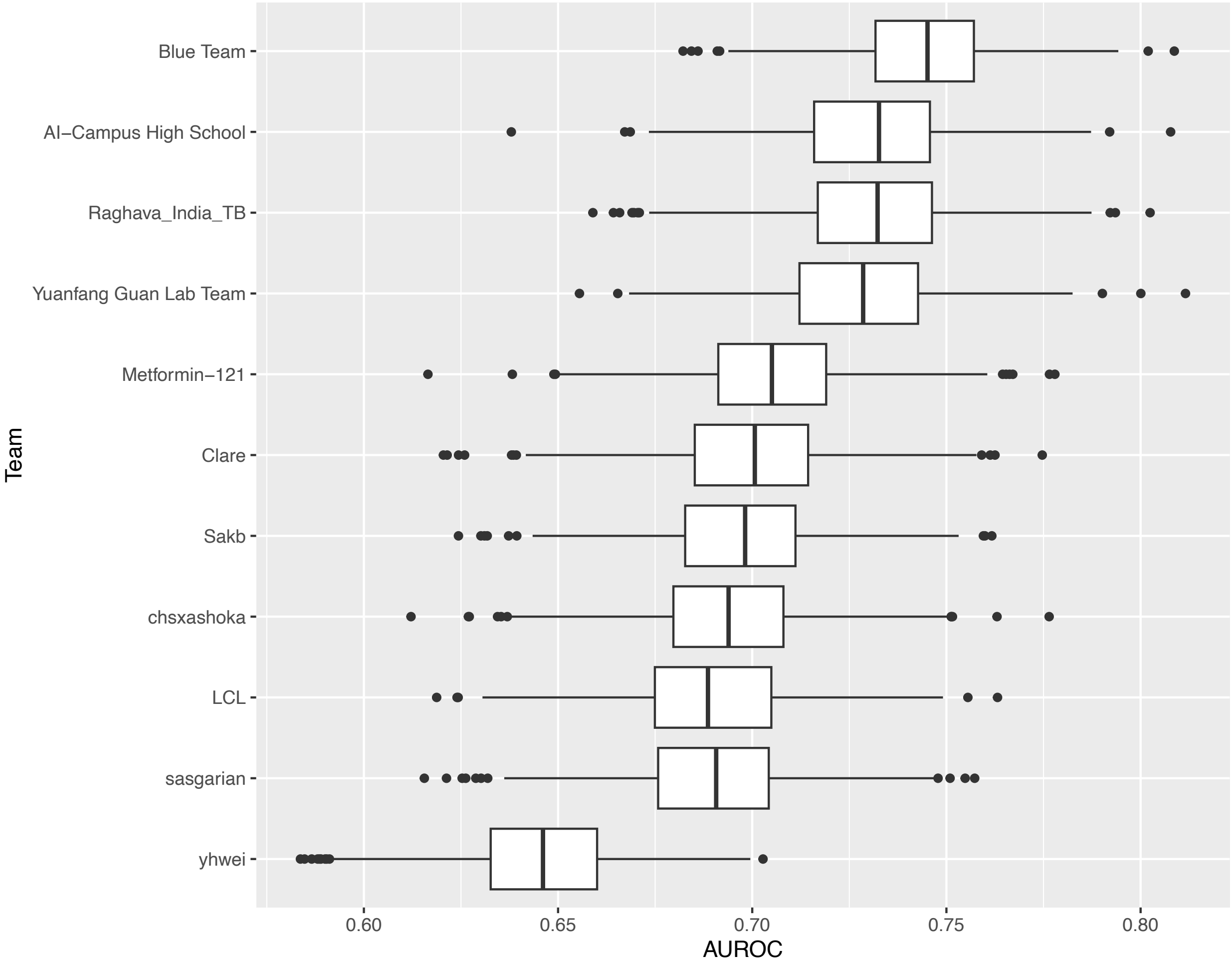

**Supplemental Figure 3. Comparison of (A) AUROC and (B) pAUROC of submissions for sub-challenge 2 (cough and clinical data) models**. Box plot of the median area under the ROC curve (AUROC) of each team with interquartile range (IQR) after 1,000 bootstraps**.** Higher pAUROC indicates greater area that captures the minimum target sensitivity (≥80%) and specificity (≥60%). For comparison, the clinical data only model is indicated in red with vertical red dashed line at the median.

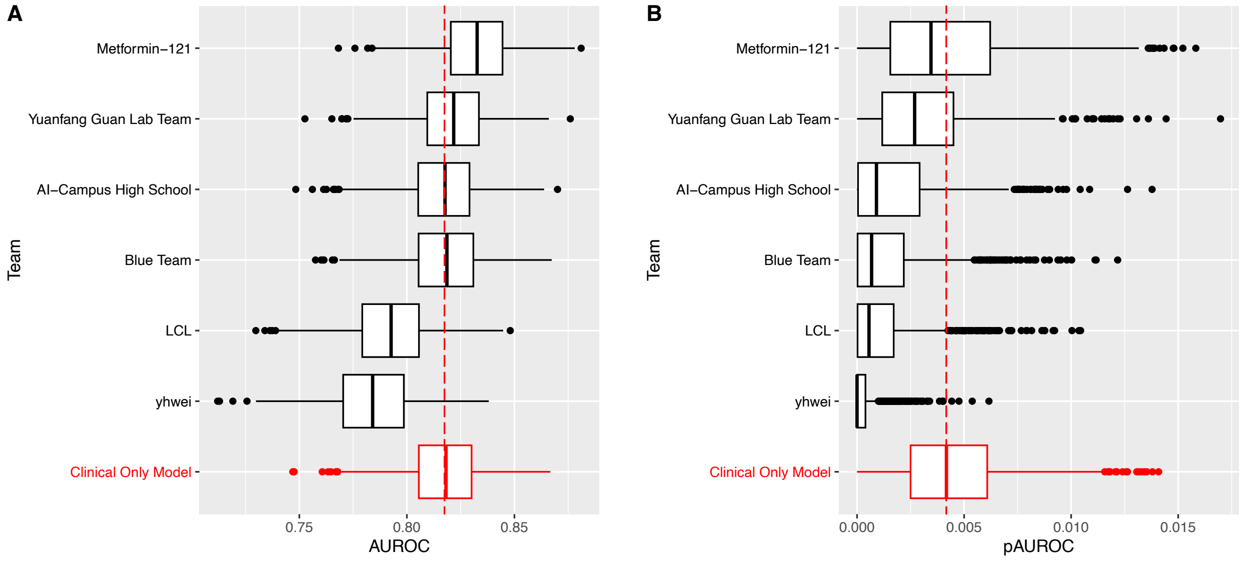

**Supplemental Figure 4. Comparison of AUROCs by country in sub-challenge 1.** Median area under the curve (AUROC) with interquartile range (IQR) based on all submissions, and stratified by country. Median AUROC indicated at the top, and winning model AUROC shown in red. SA: South Africa; MG: Madagascar; IN: India; TZ: Tanzania; UG: Uganda; PH: Philippines; VN: Vietnam.

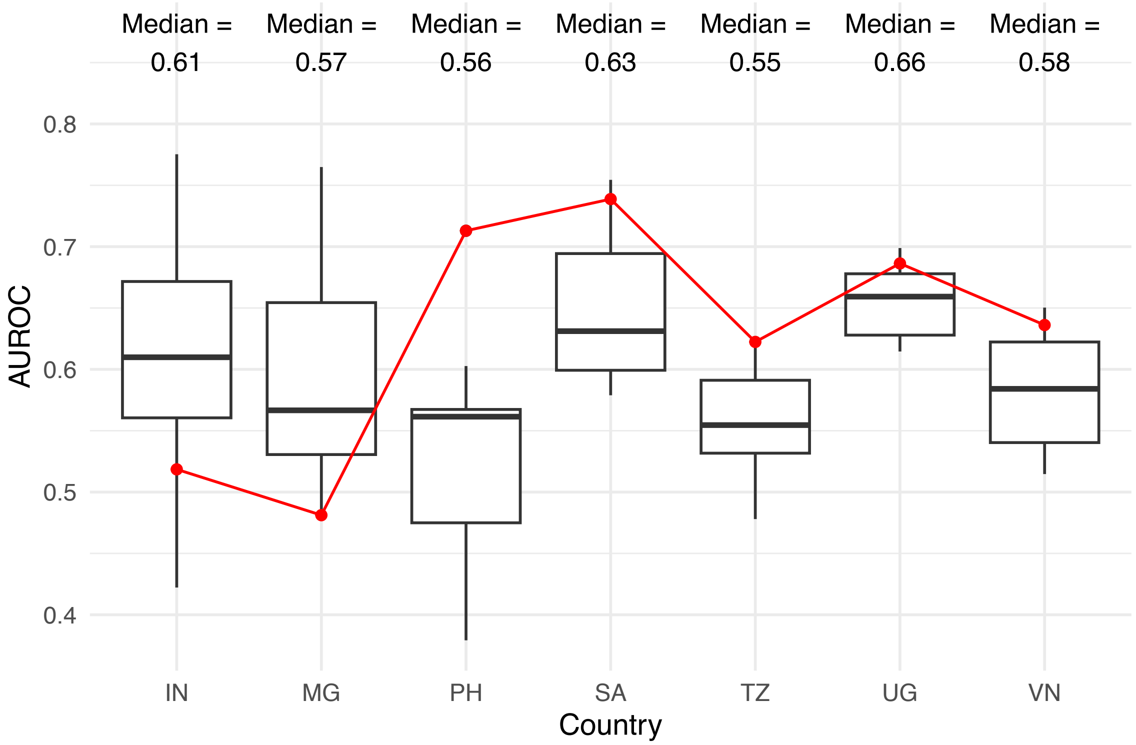

**Supplemental Figure 5. Comparison of AUROCs by sex in sub-challenge 1.** Box plot of the median area under the ROC curve (AUROC) with interquartile range (IQR) based on all submissions.

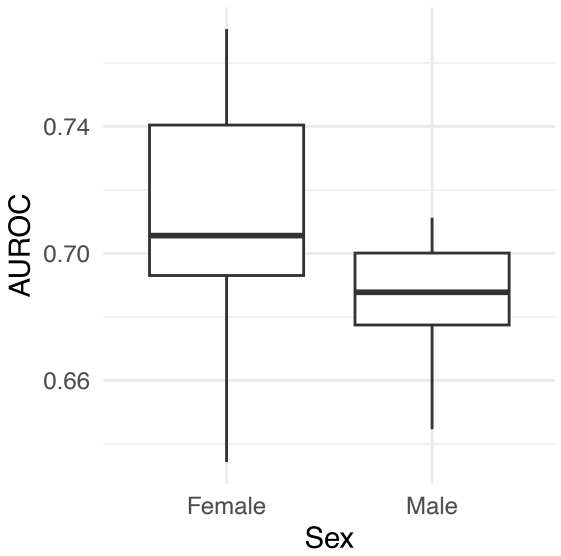

**Supplemental Figure 6.Comparison of AUROC by HIV status in sub-challenge 1.** Box plot of the median area under the ROC curve (AUROC) with interquartile range (IQR) based on all submissions.

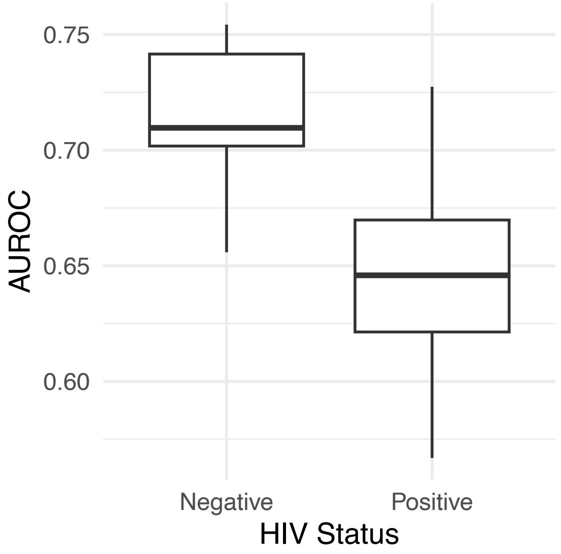

**Supplemental Figure 7. TB probability scores of the cough sound only models stratified by Xpert semi-quantitative status.** Box plot of the median probability with interquartile range (IQR). Rank indicates the final challenge ranking. Higher probability scores indicate higher likelihood that the model would classify the individual as having TB.

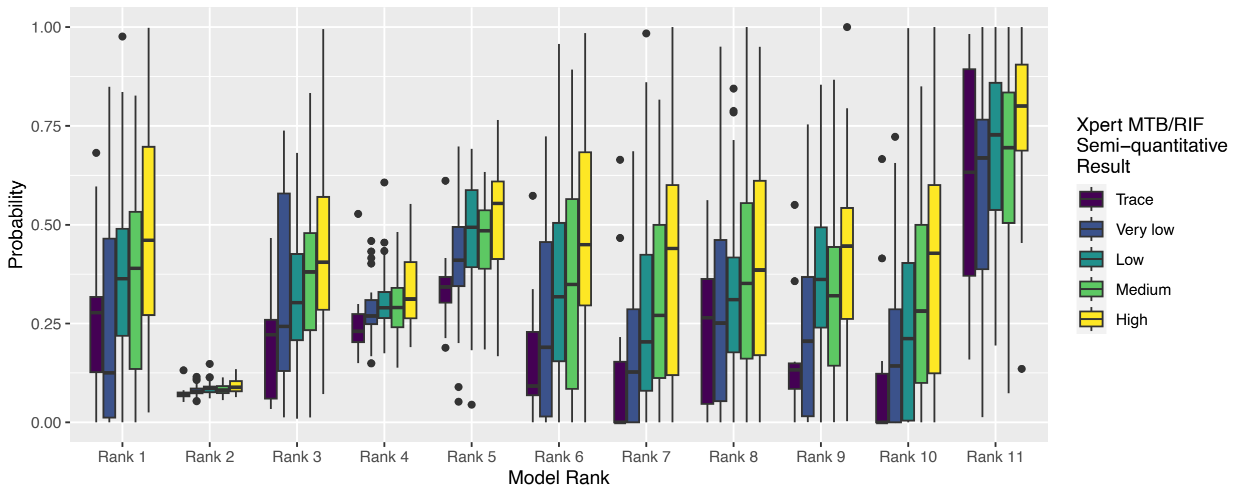
